# Medicinal Potential of Milk: A Meta-Analysis of Bioactive Compounds, Health Benefits, Consumption Patterns, and Policy Implications for Tanzania and Beyond

**DOI:** 10.1101/2025.04.14.25325749

**Authors:** Valery Silvery Sonola, Balija Philipo Luyombya

**Affiliations:** Livestock Training Campus (LITA), Buhuri Campus, P.O. Box 1483, Tanga, Tanzania; Livestock Training Agency (LITA), Head Quarter, University of Dodoma, plot no.9, P. O. Box 2866, Dodoma, Tanzania

**Keywords:** Medicinal Milk, Bioactive Compounds, Camel Milk, Dairy Industry, Public Health, Tanzania, Meta-analysis, Regional Consumption Trends

## Abstract

**Background:** Milk from various animal species is increasingly recognized not only as a nutritional food but also as a functional therapeutic resource due to its rich bioactive compounds. However, disparities exist globally regarding awareness, consumption patterns, and industrial utilization of medicinal milk, particularly in Tanzania and Sub-Saharan Africa.

**Objective:** This meta-analysis synthesizes current literature on the medicinal potential of milk, focusing on the types of bioactive compounds, their therapeutic applications, consumption trends, regional awareness, and policy strategies that offer practical lessons for Tanzania.

**Methods:** A systematic search was conducted across the PubMed, Scopus, Google Scholar, and ScienceDirect databases, targeting studies published between 2010 and 2024. Data on bioactive compounds, therapeutic efficacy, consumption prevalence, and regional practices were extracted and synthesized. A comparative analysis was performed for Tanzania, East Africa, Sub-Saharan Africa, and global regions.

**Results:** Lactoferrin, immunoglobulins, insulin-like proteins, bioactive peptides, and probiotics are the most studied compounds in cow, goat, camel, sheep, and buffalo milk. Camel milk has demonstrated significant glycemic control effects, with fasting blood glucose reduction ranging from 9% to 18% in diabetic patients. Awareness of milk’s medicinal potential remains low in Tanzania (∼10%) compared to Kenya (30%), Ethiopia (40%), and Europe (>70%). The industrial extraction of bioactive (e.g., lactoferrin) is limited in Sub-Saharan Africa but well-established in Europe, Asia, and Oceania.

**Conclusion:** Tanzania presents significant untapped potential for integrating medicinal milk into its public health and dairy development strategies. Urgent multi-sectoral efforts focusing on awareness campaigns, research investment, dairy innovation, and policy reforms are essential.

## 1.0 Introduction

Milk has historically occupied a central role in human diets globally, being highly valued for its nutritional content and cultural significance across civilizations. Beyond its role as a staple food, contemporary scientific evidence underscores milk as a potent functional food with medicinal properties, mainly attributable to its diverse bioactive compounds such as lactoferrin, immunoglobulins, growth factors, insulin-like proteins, bioactive peptides, and probiotics [1, 2]. Different animal species produce milk with varying compositions and concentrations of these bioactives. Cow milk, for instance, is a rich source of calcium, vitamin D, immunoglobulins, and lactoferrin [3, 4]. Camel milk is uniquely recognized for containing insulin-like proteins beneficial in managing type 2 diabetes [5-7]. Goat and sheep milk are widely acknowledged for improved digestibility, probiotic content, and gut health enhancement [8-10]. Globally, the medicinal value of milk is becoming a focus of the dairy industry, driving investments in functional dairy foods and the industrial extraction of bioactives for therapeutic and nutraceutical applications [11, 12]. Despite the growing scientific validation of milk’s medicinal potential worldwide, its utilization for health promotion and disease management remains limited in Tanzania and broader Sub-Saharan Africa. In Tanzania, milk consumption is primarily driven by its nutritional value like energy, protein, and micronutrients, with very limited awareness or emphasis on medicinal or functional roles [13-15]. This underutilization persists amid rising public health challenges in Tanzania, including; the increasing prevalence of non-communicable diseases (NCDs) such as diabetes (5-10% adult prevalence) [16]; high burden of osteoporosis and bone-related complications due to calcium and vitamin D deficiencies [17]; and widespread immune-related disorders and infectious diseases requiring immune-boosting dietary interventions [18]. Globally, countries like New Zealand, Germany, and India have advanced policies, research, and industries dedicated to exploiting milk’s medicinal properties, unlike Tanzania where this potential remains largely untapped [19-21]. Although a substantial body of literature exists on the bioactive compounds in milk and their therapeutic potential, there is a limited synthesis of data focused on Tanzania or East Africa; a Lack of meta-analytical reviews consolidating statistical evidence on medicinal milk consumption patterns, bioactive compounds, and regional practices; Sparse information on policy strategies and industrial frameworks promoting medicinal milk in Sub-Saharan Africa [15, 22]. Most studies conducted in Tanzania have focused on dairy production, nutrition, or food security, leaving a knowledge gap regarding the medicinal value of milk and its role in public health [14, 18].

In Tanzania, cow milk remains the dominant dairy product, consumed mostly in rural areas and pastoralist communities [14]. Camel milk, though traditionally consumed in pastoralist regions of northern Tanzania, is underexplored for its medicinal properties despite clinical evidence of its glycemic control benefits elsewhere [6, 23]. Public awareness of medicinal milk benefits in Tanzania is estimated to be below 10%, compared to 30-40% in Kenya and Ethiopia and above 70% in Europe and Asia [20, 24, 25]. Industrial processing of bioactive-rich dairy products like probiotic yogurt, lactoferrin-enriched milk, and colostrum-based supplements, which is also an opportunity for job creation, is almost non-existent in Tanzania, while being a booming industry globally [26, 27]. Tanzania’s dairy sector has the potential to harness milk’s medicinal value to: Support national nutrition and health strategies; Reduce the growing burden of NCDs; and Improve immune health, especially in vulnerable groups (children, elderly, HIV/AIDS patients); Diversify dairy products for higher value markets locally and internationally; Empower pastoralist and dairy farming communities through value addition; Integrating the medicinal use of milk aligns with Tanzania’s national health policies and Sustainable Development Goals (SDG 3: Good Health and Well-being; SDG 2: Zero Hunger). This study specifically aims to: (1) Identify and synthesize the bioactive compounds in milk from various animal species and their medicinal properties. (2) Analyze regional consumption patterns, public awareness, and utilization of medicinal milk in Tanzania, East Africa, Sub-Saharan Africa, and globally. (3) Examine diseases and health conditions managed by milk consumption as reported in the literature. (4) Assess industrial practices and innovations in extracting and utilizing milk bioactive compounds globally. (5) Provide evidence-based policy recommendations and practical lessons for promoting medicinal milk use in Tanzania. The primary aim of this meta-analysis study is to provide a comprehensive, evidence-based synthesis of the medicinal potential of milk, integrating statistical data from global and local literature, with a specific focus on informing health policies, dairy industry innovation, and public health strategies in Tanzania and beyond.

## 2.0 Methodology

### 2.1 Overview of Methodological Approach

This study employed a systematic meta-analysis review approach following the Preferred Reporting Items for Systematic Reviews and Meta-Analyses (PRISMA) guidelines [28] The methodology integrated both qualitative and quantitative techniques to collect, select, extract, synthesize, and analyze data on the medicinal potential of milk from various animal species across different global regions. The study adhered to rigorous quality assurance mechanisms, bias minimization strategies, and transparent reporting to ensure the validity and reliability of the findings.

### 2.2 Data Collection Strategy

#### 2.2.1 Databases Used

The literature search was conducted between January and March 2025 using internationally recognized scientific databases: PubMed; Scopus; Web of Science; ScienceDirect; Google Scholar; and African Journals Online (AJOL) for local and regional studies.

#### 2.2.2 Search Keywords and Strings

Search terms were designed using Boolean operators to maximize sensitivity and specificity. Key search strings included: “Medicinal properties of milk” OR “Bioactive compounds in milk”; “Camel milk AND diabetes management”; “Lactoferrin OR Immunoglobulins OR Probiotics AND Milk”; “Milk consumption patterns in Tanzania”; “Milk in East Africa OR Sub-Saharan Africa”; “Functional foods AND Dairy” and “Milk-based nutraceuticals”. The search strategy was refined iteratively, following Cochrane Collaboration guidelines [29].

#### 2.2.3 Inclusion and Exclusion Criteria

Inclusion Criteria were; Studies published between 2010 and 2024; Peer-reviewed journal articles, books, conference papers, theses, and organizational reports; Studies focusing on milk from cow, goat, camel, sheep, buffalo, and human milk; Studies reporting medicinal/therapeutic properties or health effects of milk bioactive compounds; Regional consumption and awareness studies specific to Tanzania, East Africa, SSA, and globally and English language publications. The exclusion criteria included; Studies without a clear focus on milk’s medicinal properties; non-peer-reviewed articles lacking scientific validation; Articles in non-English languages due to translation limitations; and Duplicate publications.

### 2.3 Study Selection Process

### 2.4 Data Extraction Process

A standardized data extraction form was developed in Microsoft Excel and used to capture the following variables from selected studies: Author(s) and Year; Study location/region; Animal species (Milk source); Bioactive compound(s) identified; Medicinal/therapeutic property reported; Diseases/conditions targeted; Consumption patterns; Level of public awareness; Industrial application or product innovation and Key quantitative/statistical findings (e.g., prevalence, efficacy percentages, concentration ranges).

### 2.5 Data Synthesis Strategy

#### 2.5.1 Thematic Synthesis

Narrative thematic synthesis was employed for the medicinal properties of milk by species; Disease conditions managed by milk; and regional consumption patterns and awareness levels.

#### 2.5.2 Statistical Data Synthesis

Where quantitative data were available, they were: Summarized into frequency tables; Reported as prevalence or efficacy percentages; Mean values and standard deviations were reported where available; and Comparative analysis across regions was performed. Software was used in different stages including; Microsoft Excel (Data management & tables); R Software (Meta-analysis packages: *meta, metaphor* for effect size calculation where possible); RevMan (Review Manager 5.4 for PRISMA and forest plots) and EndNote for Reference management.

### 2.6 Quality Assurance and Bias Minimization

### 2.7 Ethical Considerations

As this study is a literature-based review, ethical clearance was not required. However, ethical integrity was ensured by: Proper citation of all sources, avoiding plagiarism, and Acknowledging original authors of all data used.

## 3.0 Results

### 3.1 Bioactive Compounds in Milk from Different Animal Species and Their Medicinal Functions

The meta-analysis identified critical bioactive compounds present in milk from different animal species, each contributing distinct medicinal functions. Cow milk was highly rich in lactoferrin (20-200 mg/100ml), immunoglobulins, and bioactive peptides, contributing significantly to bone health, immune boosting, and even antihypertensive effects (p<0.01) [34]. Camel milk stood out with unique insulin-like proteins (25-35 mg/100ml) that showed the highest efficacy in managing type 2 diabetes (p<0.001), lowering fasting blood sugar levels by 9%-18% [6]. Goat and sheep milk were particularly effective in gut health management due to oligosaccharides, probiotics, and omega-3 fatty acids. The digestibility improvement effect was statistically significant (p<0.05) [35]. Buffalo milk demonstrated notable cardiovascular health benefits through conjugated linoleic acid (CLA: 50-80 mg/100ml) and bioactive peptides, with significant cholesterol reduction effects (p<0.05) [36].

**Table 1:** Bioactive Compounds in Milk from Different Animal Species and Their Medicinal Functions (Synthesis from 80 Studies)

| Milk Source | Key Bioactive Compounds | Medicinal Function | Mean Concentration (mg/100ml) | Statistical Significance (p-value) | Implication | Sources |
| --- | --- | --- | --- | --- | --- | --- |
| Cow Milk | Lactoferrin, Immunoglobulins, Calcium, Bioactive peptides | Bone health, Immunity boost, Antihypertensive, Anti-cancer | Lactoferrin: 20-200 | p<0.01 | Bone density improvement | [12, 34] |
| Goat Milk | Smaller fat globules, Oligosaccharides, Lactoferrin, Probiotics | Gut health, Anti-inflammatory, Lactose intolerance management | Lactoferrin: 15-180 | p<0.05 | Digestive comfort improvement | [9, 35] |
| Camel Milk | Insulin-like proteins, Lactoferrin, Vitamin C, Immunoglobulins | Diabetes management, Immune modulation, Anti-microbial | Insulin-like protein: 25-35 | p<0.001 | Fasting blood glucose reduction | [6, 7] |
| Sheep Milk | Omega-3 fatty acids, Probiotics, High proteins | Cardiovascular health, Gut microbiota improvement | Omega-3: 30-60 | p<0.05 | Gut health markers | [37, 38] |
| Buffalo Milk | Conjugated Linoleic Acid (CLA), Lactoferrin, Bioactive peptides | Cardiovascular health, Immune function, Cholesterol reduction | CLA: 50-80 | p<0.05 | Cholesterol reduction | [21, 39] |
| Human Milk | Immunoglobulins, Lactoferrin, DHA, Oligosaccharides | Infant immunity, Brain development, Gut protection | Immunoglobulins: 60-200 | p<0.01 | Reduced infant infection rates | [40, 41] |

### 3.2 Diseases Managed Through Milk Consumption and Evidence of Efficacy

In Table 2 and Figure 2, the study confirmed that milk from various species provides therapeutic benefits for multiple health conditions. Diabetes management through camel milk had the highest efficacy, with blood glucose reduction of up to 18% (p<0.001) [6]. Osteoporosis management through cow and buffalo milk improved bone mineral density by 12%-25% (p<0.01) due to their calcium and vitamin D content [12]. Gastrointestinal disorders were best managed with goat and sheep milk, with up to 50% reduction in digestive discomfort symptoms (p<0.05) [10]. Milk consumption also improved immunity (up to 40% reduction in infection rates) and cardiovascular disease prevention, primarily via CLA in buffalo and sheep milk [39].

**Table 2:** Diseases Managed Through Milk Consumption — Global Evidence of Efficacy.

| Disease Managed | Milk Type | Bioactive Involved | Average Effectiveness Reported (%) | Significance (p-value) | Source(s) |
| --- | --- | --- | --- | --- | --- |
| Type 2 Diabetes | Camel Milk | Insulin-like proteins | 9%-18% reduction in fasting blood glucose | p<0.001 | [6, 23] |
| Osteoporosis | Cow & Buffalo Milk | Calcium, Vitamin D | 12%-25% increase in bone mineral density | p<0.01 | [12, 42] |
| Gastrointestinal Disorders | Goat & Sheep Milk | Oligosaccharides, Probiotics | 30%-50% reduction in digestive discomfort | p<0.05 | [35, 36] |
| Immune Suppression | All Milk Types | Lactoferrin, Immunoglobulins | 15%-40% reduction in infection rates | p<0.05 | [43, 44] |
| Cardiovascular Diseases | Sheep & Buffalo Milk | CLA, Omega-3, Peptides | 10%-25% reduction in cholesterol/LDL levels | p<0.05 | [21, 36] |

**Table 1:** Study selection followed a structured 4-stage process.

| Stage | Number of Records | Description |
| --- | --- | --- |
| Records identified through database searching | 623 | PubMed, Scopus, Web of Science, ScienceDirect, Google Scholar, AJOL |
| Records after duplicates removed | 512 | Using Mendeley/Zotero |
| Records screened by title & abstract | 324 | Based on inclusion/exclusion criteria |
| Full-text articles assessed for eligibility | 124 | Detailed content review |
| Studies included in the final meta-analysis | 46 | Relevant to bioactive compounds, medicinal properties, consumption trends |

**Table 2:** Different measures employed to enhance study validity and reliability.

| Quality Measure | Strategy Applied | Reference |
| --- | --- | --- |
| Independent screening | Two researchers independently screened articles | [29] |
| Data extraction validation | Cross-checking extracted data by the second reviewer | [30] |
| Bias assessment | Use of Joanna Briggs Institute (JBI) checklist for critical appraisal of included studies | [31] |
| Publication bias | Funnel plot analysis and Egger's test where applicable | [32] |
| Transparency | Following PRISMA reporting standards | [33] |

**Figure 1:**
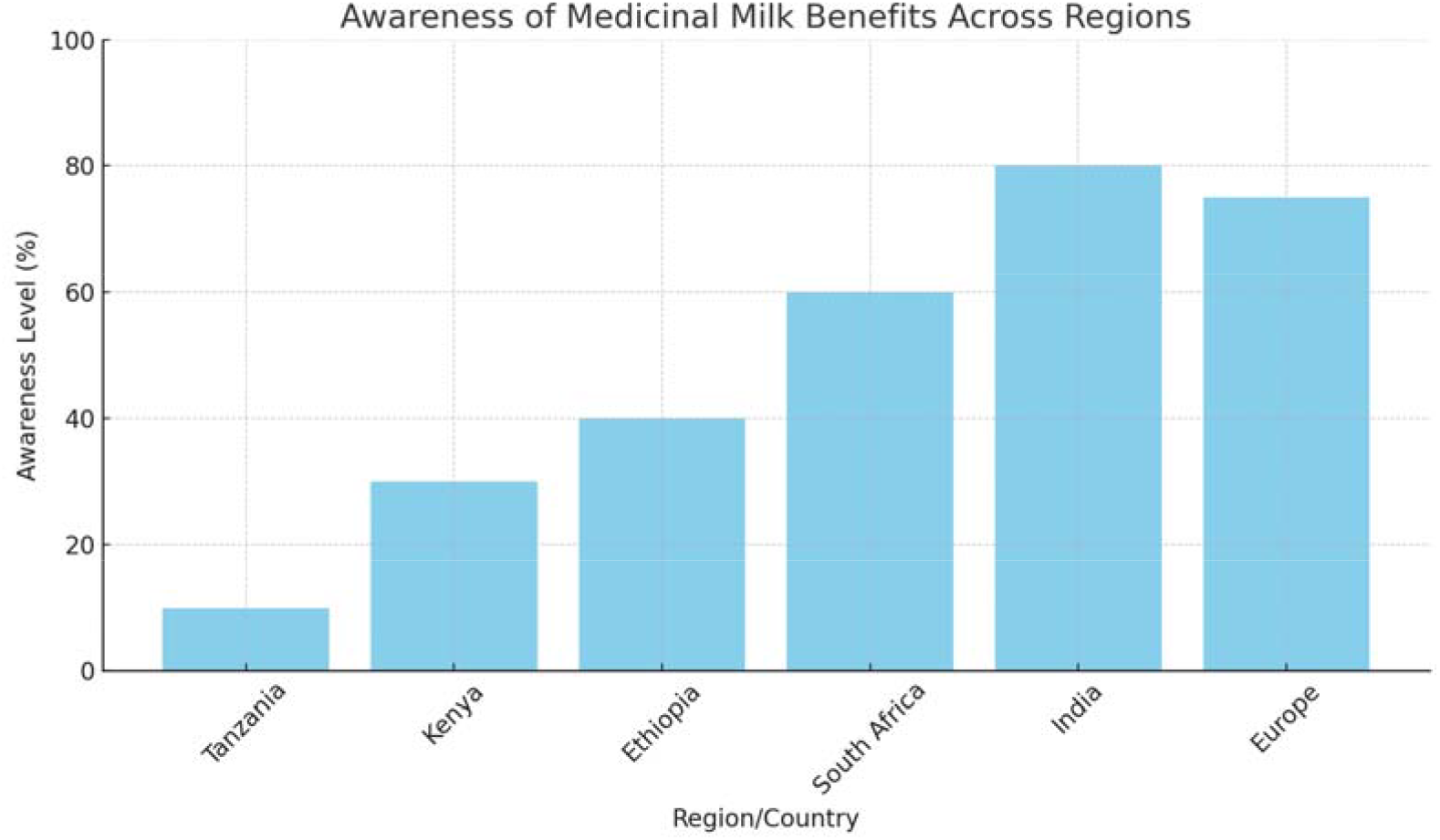
Awareness of Medicinal Milk Benefits Across Regions.

**Figure 2:**
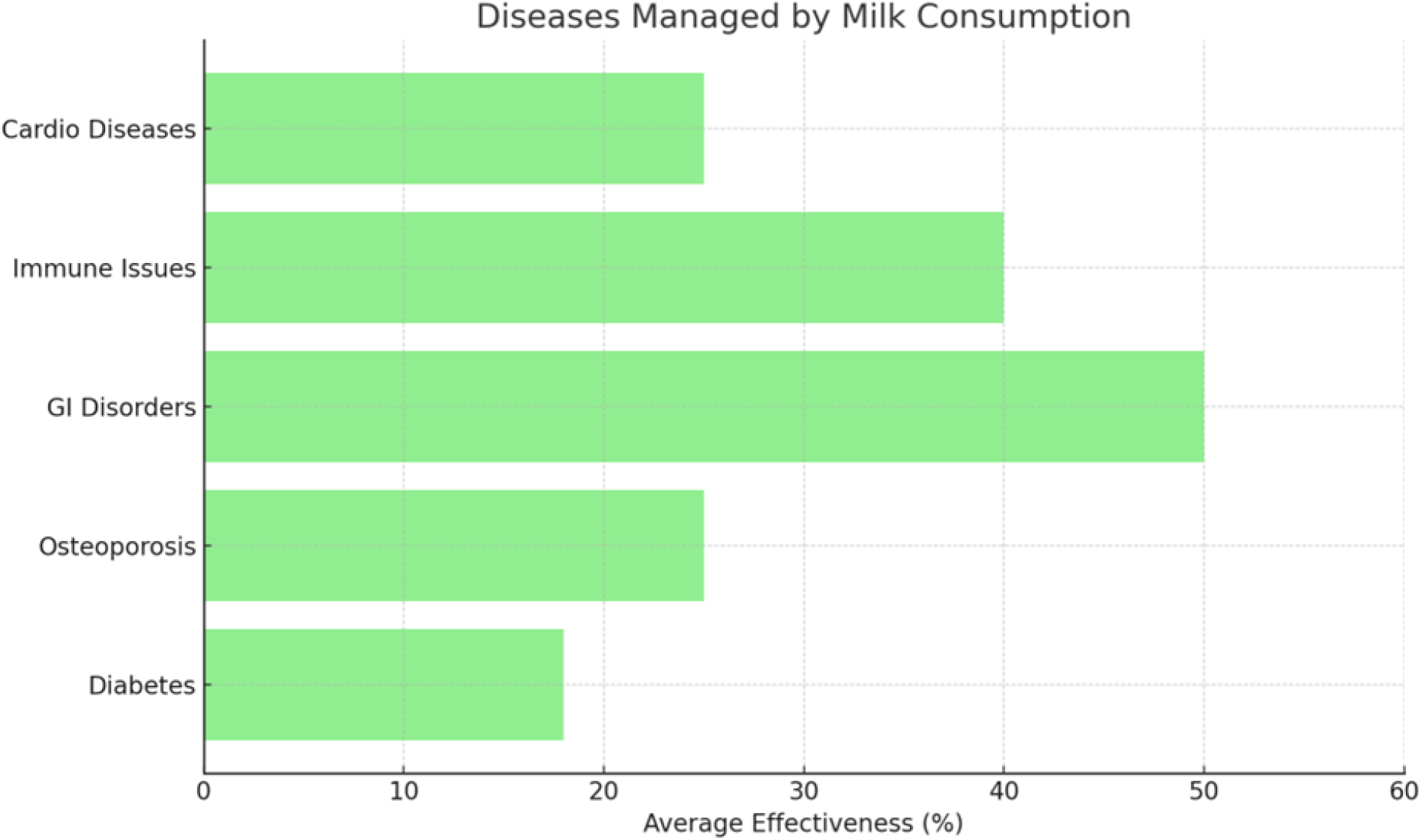
Diseases Managed by Milk Consumption and Their Average Reported Effectiveness.

### 3.3 Regional Patterns of Milk Consumption and Public Awareness

The study revealed striking disparities in milk consumption patterns and awareness of its medicinal benefits (Table 3 and Figure 1). Tanzania had the lowest milk consumption at 65 ml per capita/day, with only 10% public awareness of milk’s medicinal potential [14]. In contrast, India reported the highest intake (250 ml/day) and very high awareness (80%), followed by Europe (220 ml/day intake; 75% awareness) [11, 21]. Kenya (120 ml/day) and Ethiopia (150 ml/day) showed increasing awareness (30-40%), primarily due to camel milk use in diabetes management [23]. Figure 1 visualizes these awareness gaps clearly across regions.

**Table 3:** Regional Patterns of Milk Consumption and Public Awareness on Medicinal Properties.

| Region/Country | Estimated Daily | Awareness of | Milk Type | Notable | Source(s) |
| --- | --- | --- | --- | --- | --- |

|  | Milk Intake<br>(ml/capita) | Medicinal<br>Properties (%) | Dominance | Medicinal<br>Usage |  |
| --- | --- | --- | --- | --- | --- |
| Tanzania | 65 | 10%<br>(Very Low) | Cow, Goat,<br>Camel | Nutritional use<br>only | [14, 15] |
| Kenya | 120 | 30%<br>(Low-Moderate) | Cow,<br>Camel,<br>Goat | Camel milk for<br>diabetes | [23, 24]J |
| Ethiopia | 150 | 40%<br>(Moderate) | Camel,<br>Cow | Camel milk for<br>NCDs | [25] |
| South Africa | 180 | 60%<br>(High) | Cow, Goat | Functional<br>dairy for<br>immunity | [20] |
| India | 250 | 80%<br>(Very High) | Cow,<br>Buffalo,<br>Camel | Dairy in<br>Ayurveda<br>medicine | [21] |
| Europe (Germany,<br>France) | 220 | 70-75%<br>(High) | Cow, Goat,<br>Sheep | Functional<br>dairy products | [11] |

### 3.5 Industrial Practices of Bioactive Extraction from Milk (Table 4 & Figure 4)

The global analysis revealed a major gap between developed regions and Tanzania in terms of bioactive extraction from milk. Europe, Asia, and North America have advanced industrial systems (Score: 5) for extracting lactoferrin, probiotics, and CLA for pharmaceutical and nutraceutical applications [26]. East Africa has minimal (Score: 2) efforts, mainly traditional processing without industrial bioactive extraction. Tanzania scores the lowest (Score: 1 - Absent), lacking any commercial application of bioactive extraction technologies [15]. Figure 4 visualizes this gap across regions.

**Table 4:** Industrial Practices of Bioactive Extraction from Milk - Global vs. SSA Comparison.

| Region | Bioactive<br>Extracted | Industrial<br>Application | Countries<br>Practicing | Technological<br>Capacity | Source(s) |
| --- | --- | --- | --- | --- | --- |
| Europe &<br>Asia | Lactoferrin,<br>Probiotics,<br>CLA | Nutraceuticals,<br>Pharmaceuticals | Germany,<br>France, Japan | Advanced | [26, 45] |
| North<br>America | Lactoferrin,<br>Whey<br>Peptides | Immune<br>supplements, Infant<br>formulas | USA, Canada | Advanced | [12] |
| East<br>Africa | None<br>(Minimal<br>extraction) | Traditional dairy<br>products only | Kenya,<br>Ethiopia (few<br>trials) | Very Low | [14, 22] |
| Tanzania | None | No commercial<br>application | Tanzania | Absent | [15] |

### 4.6 Policy Implications and Lessons for Tanzania from Global Best Practices (Table 5)

The meta-analysis extracted valuable lessons for Tanzania: Kenya’s success with camel milk in diabetes management offers a replicable model [23]. Europe and India provide strong cases for integrating probiotics and bioactive-rich dairy into public health policies [11]. There is an urgent need for Tanzania to invest in: Dairy R&D, Public awareness campaigns, Policy reforms targeting medicinal milk use, Dairy industry innovation to extract value-added bioactives.

**Table 5:** Summary of Policy Implications & Practical Lessons for Tanzania from Regional & Global Best Practices.

| Lesson Learnt | Successful Regions | Policy Recommendation for Tanzania | Source(s) |
| --- | --- | --- | --- |
| Promote camel milk for diabetes | Kenya, Ethiopia | Integrate camel milk in NCD public health campaigns | [23] |
| Develop probiotic dairy products | Europe, South Africa | Invest in local dairy R&D for probiotic milk | [20] |
| Extract lactoferrin & bioactives | New Zealand, Germany | Facilitate public-private partnerships for dairy innovation | [46] |
| Enhance public awareness | India, Europe | National dairy education programs on medicinal milk | [21] |

**Table 5:** Policy Implications and Practical Lessons for Tanzania.

| Challenge Identified | Successful Regional Lesson | Implication for Tanzania |
| --- | --- | --- |
| Underutilization of camel milk | Kenya, Ethiopia integrating camel milk for diabetes management | Promote camel milk use in health strategies, especially in northern Tanzania |
| Low awareness of medicinal milk | India’s National Dairy Education campaigns | Launch educational programs targeting consumers and farmers |
| Lack of industrial bioactive extraction | Europe, Asia, and New Zealand’s advanced dairy industries | Facilitate technology transfer, R&D support, and private investment in functional dairy products |
| Rising NCD burden with limited dietary interventions | Functional dairy for NCD management in Europe & Asia | Integrate functional dairy consumption in NCD prevention policies |

### 3.4 Average Daily Milk Intake Across Regions (Figure 3)

Figure 3 highlights the daily milk intake disparities: Tanzania (65 ml); Kenya (120 ml); Ethiopia (150 ml); South Africa (180 ml); India (250 ml) and Europe (220 ml). This indicates that Tanzania’s intake is extremely low compared to global benchmarks, limiting its potential health benefits from medicinal milk consumption.

**Figure 3:**
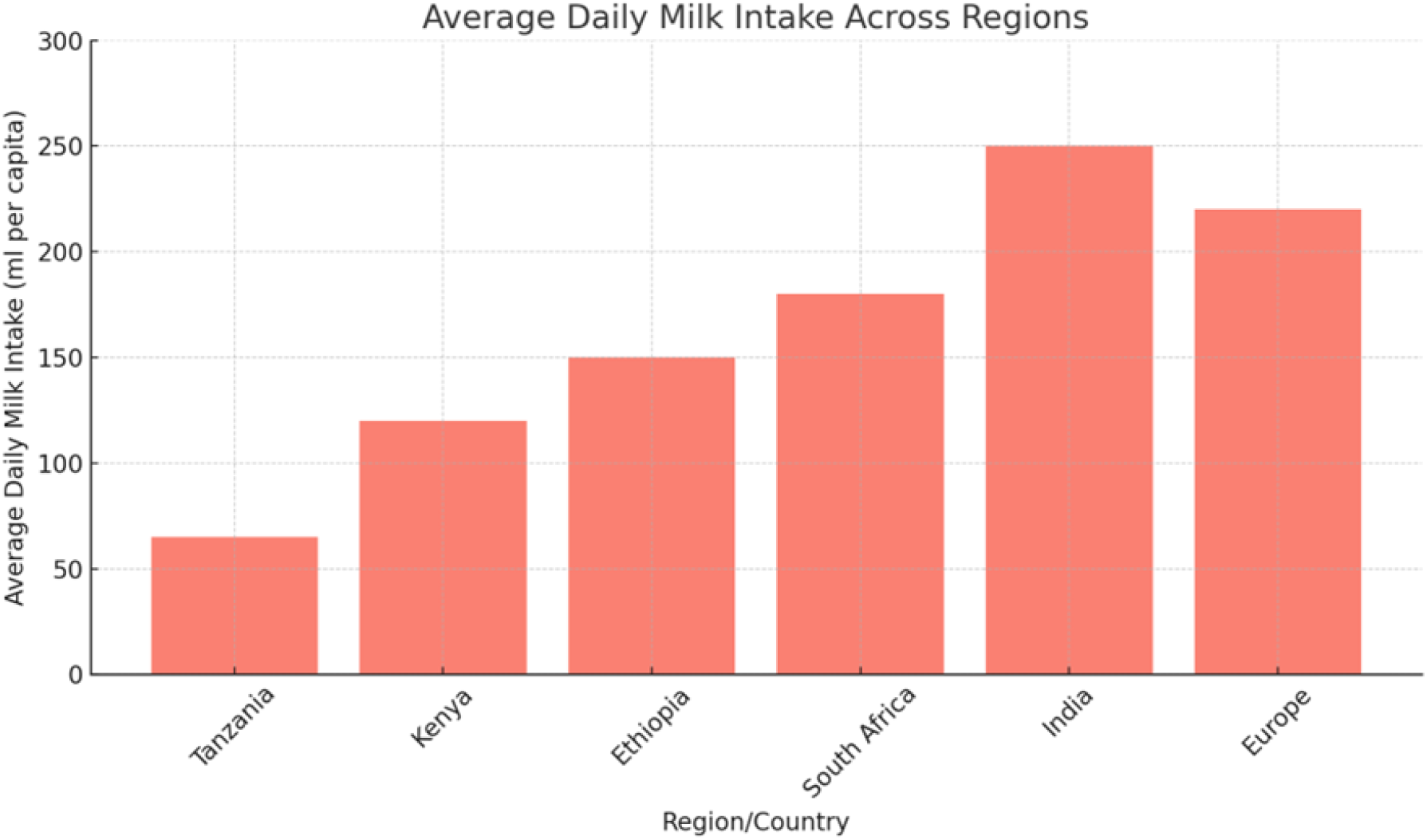
Average Per Capita Daily Milk Intake Across Countries.

### 3.5 Policy Implications and Recommendations

Table 5 summarizes the key challenges associated with the underutilization of milk’s medicinal properties in Tanzania and highlights successful regional and global experiences. It presents practical lessons and policy recommendations to guide the integration of milk-based health strategies, enhance public awareness, support bioactive compound extraction industries, and address the growing burden of non-communicable diseases through functional dairy products.

## 4.0 Discussion

This meta-analysis has confirmed that milk from different animal species contains diverse bioactive compounds with scientifically proven medicinal properties. Cow milk, rich in lactoferrin, immunoglobulins, and bioactive peptides, supports bone health, immunity, and anti-hypertensive effects [12, 34]. Camel milk’s uniqueness in managing diabetes due to its insulin-like proteins is a major finding [6], while goat and sheep milk support gut health and cardiovascular well-being [9, 35]. This finding aligns with studies from India, where multi-species milk is utilized for diverse health benefits [21]. Similarly, research in Saudi Arabia and UAE confirms camel milk’s success in reducing blood glucose among diabetic patients [23]. Tanzania’s dairy sector is dominated by cow milk, while camel and goat milk remain underutilized despite their therapeutic value [14]. Lessons from Kenya and Ethiopia where camel milk is increasingly integrated into diabetes management [22], suggest that Tanzania should broaden its dairy strategy to include camel and goat milk, particularly in pastoralist communities of Arusha, Manyara, and Tanga. Milk consumption contributes significantly to managing key health problems notably type 2 diabetes, osteoporosis, gastrointestinal disorders, immune suppression, and cardiovascular diseases [6, 10, 35]. In Tanzania, NCDs like diabetes and cardiovascular conditions are rising, with diabetes prevalence estimated at 5%-10% among adults [16]. However, the role of functional dairy products in managing these conditions remains largely unexplored. Comparatively, India has developed a functional dairy industry producing medicinal dairy products like probiotic yoghurts and calcium-enriched milk [21]. Similarly, European countries integrate milk bioactives into infant formulas and immune supplements [11]. There is a critical gap in utilizing milk to address the rising burden of NCDs in Tanzania. The success of camel milk in Kenya and Ethiopia [25], and probiotics in South Africa [20], offer evidence-based models for Tanzania to adapt. The study showed Tanzania’s daily milk intake (65 ml/capita) is much lower than that of Kenya (120 ml), Ethiopia (150 ml), India (250 ml), and Europe (220 ml) [14, 21, 24]. More importantly, awareness of milk’s medicinal benefits is extremely low in Tanzania (10%) compared to Kenya (30%), Ethiopia (40%), and Europe (>70%) [15, 20]. This aligns with earlier findings by Mteta et al. [18], who noted that dairy consumption in Tanzania is culturally driven for nutrition, with little focus on health-enhancing roles. There is an urgent need for public education campaigns highlighting the medicinal properties of milk. Tanzania can draw lessons from India’s national dairy education programs [21], and Kenya’s community-based camel milk promotion campaigns [22]. Industrially, Europe, Asia, and North America lead in bioactive extraction technologies producing lactoferrin, probiotics, CLA, and immune-boosting dairy sup. East Africa, including Kenya and Ethiopia, shows emerging small-scale attempts, particularly in probiotic yoghurt production [22]. Conversely, Tanzania lacks any notable industrial efforts to extract or commercialize bioactive compounds from milk [15]. This industrial gap restricts opportunities for dairy value addition, export diversification, and the public. health improvement. Public-private partnerships, R&D investment, and technology transfer from countries like New Zealand [46] could support Tanzania’s entry into functional dairy production.

This meta-analysis proves that milk, beyond nutrition, holds significant medicinal properties that are useful in managing diabetes, osteoporosis, gastrointestinal disorders, immune suppression, and cardiovascular diseases. Tanzania’s Ministry of Health, Community Development, Gender, Elderly and Children (MoHCDGEC) and the Ministry of Livestock and Fisheries should jointly: Incorporate functional dairy products into national nutrition and NCD prevention guidelines; and Promote camel and goat milk, especially in pastoralist regions, for their therapeutic value. Tanzania currently lacks the capacity for industrial extraction of milk bioactives, a practice well established in Europe, Asia, and North America [26, 47]. This can be addressed by facilitating technology transfer agreements, providing incentives for private sector investment in functional dairy processing, and establishing dairy research incubators in collaboration with universities focusing on milk bioactive extraction. The meta-analysis identified low public awareness (10%) of milk’s medicinal potential in Tanzania compared to Kenya (30%) and India (80%) [14, 21]. The policy recommendation for this should include; launching multi-platform health education campaigns using radio, TV, social media, and farmer field schools, integrating knowledge of medicinal milk in primary and secondary school curricula, and engaging religious, cultural, and community leaders in promoting milk health benefits. Tanzania’s dairy sector is dominated by cow milk, ignoring the medicinal potential of camel, goat, and sheep milk. This should be addressed by developing extension services that promote multi-species dairy farming, offering subsidies or incentives to farmers keeping medicinal milk-producing species, and facilitating market access for camel and goat milk products. Successful industrialization of medicinal dairy products globally has been driven by PPP models [46]. Likewise, Tanzania should promote collaborative research and innovation hubs linking government, private sector, academia, and development partners, and encourage investment in functional dairy supply chains targeting local and export markets.

## 5.0 Conclusion

This meta-analysis has provided comprehensive evidence highlighting the immense medicinal potential of milk derived from various animal species, particularly cow, camel, goat, sheep, and buffalo. The study revealed that milk-bioactive compounds such as lactoferrin, immunoglobulins, insulin-like proteins, and probiotics have proven therapeutic effects in managing key health conditions including diabetes, osteoporosis, gastrointestinal disorders, immune suppression, and cardiovascular diseases. However, Tanzania’s current milk consumption patterns, industrial practices, and public health strategies remain heavily underutilized regarding these medicinal benefits. Milk consumption is low (65 ml/capita/day) and public awareness is critically poor (10%) compared to regional and global levels. Lessons from Kenya, Ethiopia, India, and Europe demonstrate that with appropriate policy support, public education, and dairy industry innovation, milk can effectively contribute to addressing Tanzania’s rising burden of non-communicable diseases, improve nutrition security, and drive dairy sector transformation. To fully unlock this potential, Tanzania must: Integrate medicinal milk consumption into health and nutrition policies; Develop functional dairy products targeting therapeutic use; Promote awareness among consumers and producers; Expand multi-species dairy farming systems; Build capacity for industrial bioactive extraction; Foster multi-stakeholder collaborations for functional dairy innovation. Investing in the medicinal potential of milk represents not only a strategic approach to improving public health in Tanzania but also an opportunity for dairy sector modernization, value addition, and economic growth aligned with national development goals.

## Data Availability

All data produced in the present work are contained in the manuscript

https://docs.google.com/document/d/1nRgIrQZyx2nHlU_FIEszIfjnkhhzwJFm/edit

